# Therapeutic efficacy of four antiviral drugs in treatment of COVID-19: A protocol for systematic review and Network Meta-analysis

**DOI:** 10.1101/2022.04.05.22273473

**Authors:** Liang-kun Zhang, Wen-chao Gu, Ling Li, Chen Jian

## Abstract

**Introduction:** The COVID-19 pandemic has resulted in a global crisis in public health, however, there are still no safe and effective drugs to resist COVID-19 until now. We will use network meta-analysis to analysis available evidence from RCTs to compare the safety and efficacy of four antiviral drugs (including Ribavirin, Arbidol, Chloroquine Phosphate and Interferon) alone or in combination, in patients with COVID-19 on the basis of standard treatment, to reveal the robustness and strength of evidence for relative efficacy against COVID-19, which will provide better evidence for future clinical decision-making.

**Methods:** Using English and Chinese search strategies to search 8 databases including PubMed, Web of Science, Embase, the Cochrane Library, CNKI, CBM, WANFANG Database and VIP. In addition, manual search for references in publications has been supplemented by electronic search. To enhance the effectiveness of this study, only randomized controlled trials of four antiviral drugs (Ribavirin, Arbidol, Chloroquine Phosphate, Interferon) used alone or in combination with the primary therapy shall be included.

**Analysis:** The nucleic acid turning negative, complete absorption of lung inflammation, adverse reactions, aggravation and death shall be the primary outcome measures; whereas temperature return to normal, hospitalization, and positive rate after discharge will be the secondary outcomes. To ensure the quality of the systematic evaluation of this study, study screening, data extraction and quality evaluation will be carried out independently by two reviewers, and any differences will be resolved through consultation between them or by a third reviewer.

**Ethics and dissemination:** This systematic review will evaluate the efficacy of four antiviral drugs (Ribavirin, Arbidol, Chloroquine Phosphate and Interferon) for Covid-19 in adults. Since all included data will be obtained from published articles, it does not require ethical approval and will be published in a peer-reviewed journal.

**PROSPERO registration number:** CRD42022300104.

## Introduction

Coronavirus disease (COVID-19) is an infectious disease caused by severe acute respiratory syndrome coronavirus (SARS-CoV-2), which manifestations include asymptomatic carriers and fulminant disease characterized by sepsis and acute respiratory failure [1-3]. SARS-CoV-2 is spread primarily via respiratory droplets during close face-to-face contact[4]. Pathogenesis of COVID-19 may be included coagulopathy, endothelial dysfunction, and excessive release of pro-inflammatory cytokines[5]. COVID-19 involved multiple systems such as respiratory system, reproductive system and digestive systems[6-10], which could rapidly develop into acute respiratory distress syndrome, septic shock or multiple organ failure in severe cases[11, 12]. As of 27 February 2022, over 433 million confirmed cases and over 5.9 million deaths have been reported globally, according to date released by the WHO[13]. In the face of this pandemic, the Chinese government has provided valuable lessons for other countries in the treatment of COVID-19, and issued the latest edition of Diagnosis and Treatment Protocol for Novel Coronavirus Pneumonia (Trial 8th revised edition)[14] on April 14, 2021. Its main efficacy intervention including antiviral therapy, immunotherapy, glucocorticoid therapy, traditional Chinese medicine therapy and so on. Among which, antiviral drugs mainly include interferon (IFN), ribavirin, chloroquine phosphate and arbidol.

Ribavirin (a nucleoside analog) is a broad-spectrum antiviral drug, which is mainly used in the treatment of respiratory syncytial virus (RSV), influenza virus and adenovirus infection[15, 16]. The U.S. Food and Drug Administration and part international guidelines recommend ribavirin in combination with interferon to treat chronic hepatitis C[17-19], however, hemolytic anemia is one of the most serious adverse reactions to long-term and high-dose use of ribavirin[20, 21]. As the global COVID-19 pandemic continues, studies have shown that early use of ribavirin antiviral regimen may lead to increased efficacy[22].

Chloroquine phosphate is a derivative of chloroquine, which has particulars such as quick absorption of drug, fewer adverse reactions. The mechanism may be to interfere with endosome acidification, protect low pH-dependent targets in vivo, block angiotensin converting enzyme 2 (ACE2) glycosylation, and prevent cells from binding to viral receptors to achieving the purpose of antiviral[23-25]. In addition, it can also inhibit the production and release of inflammatory cytokine in the middle and late period of viral infection[26]. At present, there are a number of existing studies on chloroquine phosphate which could suppress severe acute respiratory syndrome virus (SARS-CoV)[27, 28], influenza virus[29], Dengue virus[30], human immune deficiency virus (HIV)[31]. Since outbreak of COVID-19, both Diagnosis and Treatment Protocol for Novel Coronavirus Pneumonia (Trial 8th revised edition)[14] and the Chinese expert consensus[32] have recommended chloroquine phosphate for treatment of COVID-19.

Arbidol is a small indole derivative with immune-enhancement and broad-spectrum antiviral effects[33, 34]. As the only hemozoin inhibitor at present, it is mainly used to prevent and treat upper respiratory tract infections caused by influenza A/B viruses[35]. Studies have found that arbidol could not only kill virus directly, inhibit virus proliferation and antigen expression, but also activate 2, 5-oligosadenylate synthase, which could specifically inhibit fusion of the virus membrane with the host cell membrane to block virus entry target cell and inhibit the replication of the virus to achieve the antiviral effect[36-38]. In addition, it could enhance humoral immunity and cellular immunity function, induce cells to produce interferon, regulate release of inflammatory cytokines and activate phagocytes, regulate CD4, CD8 levels to play an antiviral role[39, 40]. Arbidol has been shown to be effective against COVID-19 in clinical studies[41, 42], and six clinical trials have been registered.

IFN was discovered by Issacs and Lindenmann in 1957 during their study on phenomenon of virus interference[43], which are classified by type 1, type 2 and type 3. Type 1 IFN is secreted by cells infected with virus and has strong antiviral, antitumor, immunomodulatory properties; Type 2 IFN, secreted by activated T cells, is mainly involved in the immune regulation; Most cells could secrete type 3 IFN, which generate beneficial mucosal immunomodulatory effects[44-46]. Among them, type 1 and type 3 IFN are classical antiviral interferons. At present, IFN has achieved remarkable results in the treatment of COVID-19[47, 48]. The mechanism may be that the increase in post-infection of SARS-CoV-2 results in an imbalance of IFN response, leading to excessive inflammatory response and even cytokine release syndrome, inducing to multiorgan failure[49, 50].

However, most of researches only reported clinical efficacy of the four drugs in treatment of COVID-19 alone or in combination, which were single center studies with a small sample size, different degrees of blinding, and lack of comparison of clinical efficacy and safety evaluation of the four drugs. Therefore, the aim of this study is to analyze the results of randomised controlled trials (RCTs) to ascertain the efficacy and safety of the four antiviral drugs (α-IFN, ribavirin, chloroquine and arbidol) used alone or in combination for the treatment of COVID-19, which will provide reliable evidence-based clarification of the efficacy for the treatment of COVID-19 in adults.

The proposed date for the completion of this study is June 15, 2022.

## Materials and methods

The protocol of this systematic review and network meta-analysis will be reported in accordance with the guidelines of the Preferred Reporting Items for Systematic Reviews and Meta Analyses for Network Meta-Analyses (PRISMA-NMA) extension statement[51]. In January 2022, we have obtained the registration number (CRD42022300104) of this review on the platform of PROSPERO (https://www.crd.york.ac.uk/PROSPERO/#myprospero).

### Inclusion and exclusion criteria

Detailed Inclusion and exclusion criteria were developed following 5 main principles of PICOS (Participants, Interventions, Comparators, Outcomes, and Study design)[52], which were summarised in Table 1.

**Table 1.** Eligibility criteria of included studies

|  |  |
| --- | --- |
| Participants | Participants aged over 18 years with positive novel coronavirus nucleic acid test |
| Interventions | Four antiviral drugs in alone or in combination |
| Comparators | Fundamental treatment in general treatments, immunotherapy, glucocorticoid therapy, traditional Chinese medicine therapy, and so on |
| Outcomes | Primary outcome measures included nucleic acid turning negative, complete absorption of lung inflammation, adverse reactions, aggravation or death; Secondary outcomes include temperature return to normal, hospitalization, and positive rate after discharge etc. |
| Study design | RCTs with no restrictions on language |

Participants must be aged over 18 years and have a positive novel coronavirus nucleic acid test (RT-PCR reaction) who had been clearly diagnosed with COVID-19. there were no strict restrictions on gender, racial group, or any non-specified baseline characteristics. Eligible interventions in the RCTs that include at least one of the four antivirals (α-IFN, ribavirin, chloroquine and arbidol) in addition to fundamental treatment including general treatments, immunotherapy, glucocorticoid therapy, traditional Chinese medicine therapy, and so on against controls only with fundamental treatment. There are no clear restrictions on the dosage and administration of therapeutic drugs. The main primary outcome measures included nucleic acid turning negative, complete absorption of lung inflammation, adverse reactions, aggravation or death; Secondary outcomes include temperature return to normal, hospitalization, and positive rate after discharge etc. In addition to the above criteria, eligible study designs must be RCTs with English or Chinese on language. Non-RCTs, case reports, clinical experience summary, reviews, animal studies, and repeatedly publications will be excluded. Meanwhile, literatures with unclear research results and incomplete data will be also excluded.

### Data sources

Two investigators will search eight electronic databases (PubMed, Web of Science, Embase, the Cochrane Library, CNKI, CBM, WANFANG database and VIP) for RCTs of four antiviral drugs for COVID-19, starting from the inception dates of databases. References cited in the included publications will be manually searched to check for additional relevant randomised controlled trial reports. Search terms include: Ribavirin, Arbidol, Chloroquine Phosphate, Interferon, COVID-19, 2019-NCOV, SARS-COV-2. Different databases choose the corresponding combination of subject words, free words and keywords. Table 2 takes the initial retrieval strategy of PubMed database as an example, which will be adjust search keywords according to the needs of other databases.

### Selection of studies

All retrieved records will be imported into EndNote (X9) to establish an information database and duplicate references will be deleted. Data will be extracted and cross-examined independently by two investigators based on pre-established forms.

**Table 2.**
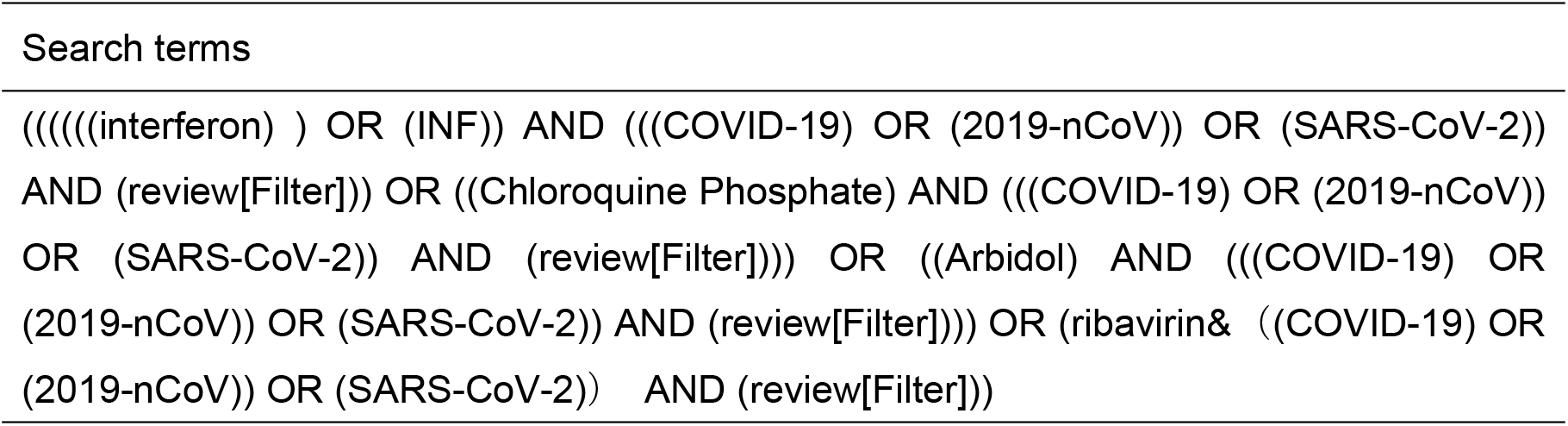
PubMed search strategies

Disagreements between investigators will be resolved by discussion with the principal investigator. The process of study selection will be summarized in the PRISMA[53, 54] flowchart in Figure 1.

**Fig 1.**
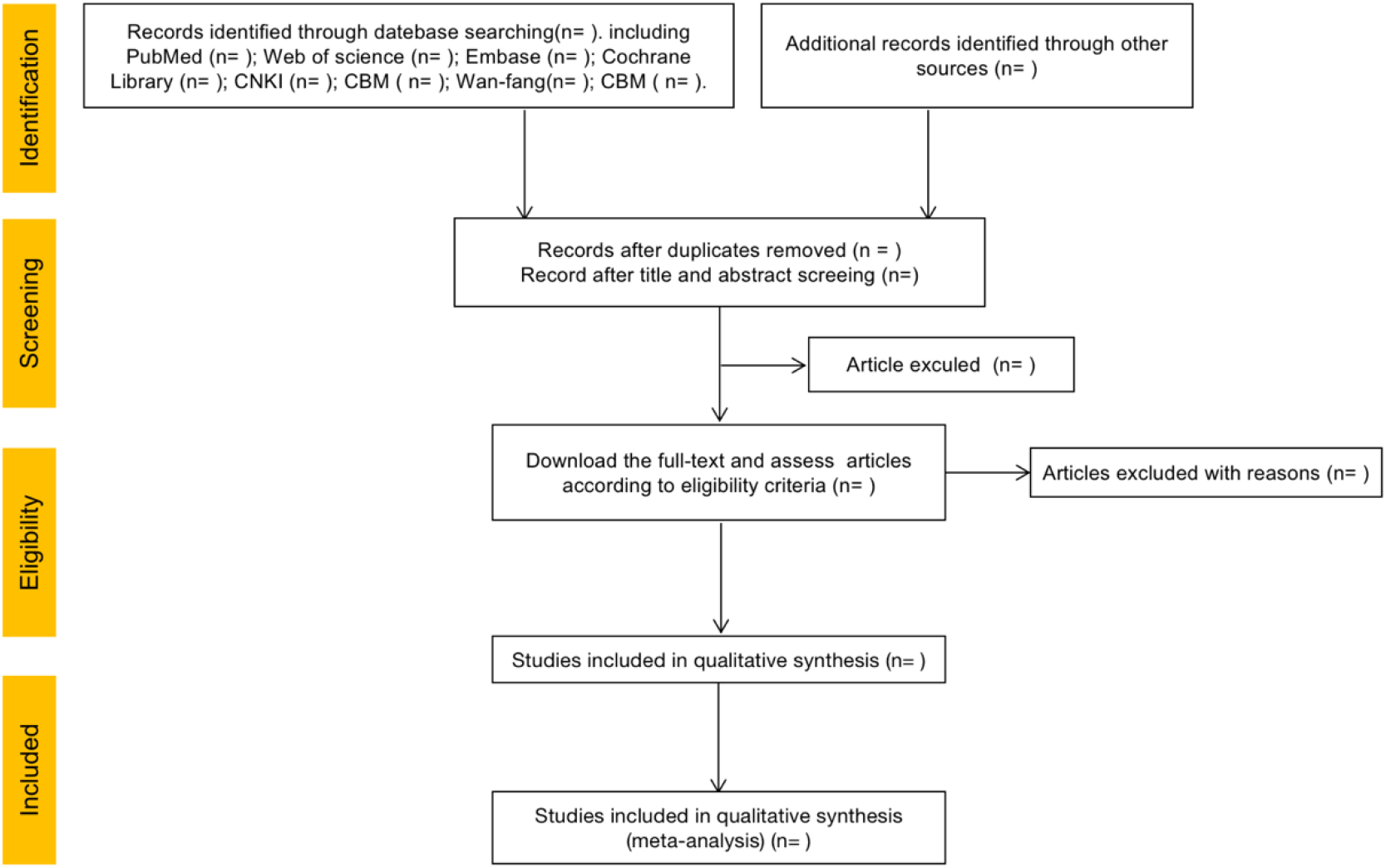
PRISMA flow diagram of study and exclusion.

### Data extraction and management

Following specific information will be extracted:

a. study characteristics include title, author names, journal of publication, year, follow-up durations, etc.
b. participant/population characteristics including sample sizes, proportions of males, age, etc.
c. comparative intervention measures including administration regimens, drug names, doses and administration time etc.
d. comparative outcomes including nucleic acid turning negative, complete absorption of inflammation, temperature recovery, rate of repositive after discharge, significant improvement of clinical symptoms, adverse reactions, and deaths etc.
e. Design types and quality evaluation information of the included literature.

The above information is ultimately cross-checked by two reviewers, and any differences may be resolved by consensus or arbitration by a third reviewer.

### Risk of bias assessment

Two investigators will independently assess all included studies according to the Cochrane tool for assessing risk of bias[55] in randomised trials described in the Cochrane Handbook for Systematic Reviews of Interventions. The risk table will assess 6 items, including selection of reported result, measurement of outcome, missing outcome data, deviations from intended interventions, randomization process and overall bias. Each item will be judged as “high risk of bias,” “some concerns,” or “low risk of bias” by the answers to the respective questions. Differences between investigators will be resolved through discussion with each other or consultation with the principal investigator.

### Meta-analysis

We will use Stata version 17 (StataCorp, College Station, TX) to select appropriate models for network meta-analysis of each outcome indicator according to its heterogeneity. Continuous outcomes will be expressed by mean difference (MD) and 95% confidence interval (CIs), while dichotomous outcomes will be expressed by ratio ratio (RR) and 95% Cis. Therapeutic measures were ranked according to SUCRA results. We will assess heterogeneity by I^2^ index[56]. If there is no significant heterogeneity (I^2^<50%) between paired studies, the fixed-effect model will be used for meta-analysis; otherwise, the random effect model will be used for evaluation. If the clinical trial data provided is quite heterogeneous, but we do not perform a meta-analysis but a descriptive analysis.

At the same time, if possible, we shall perform subgroup analyses based on study characteristics, such as duration of treatment and disease stage, to interpret heterogeneity. Sensitivity analyses will also be conducted to screen for studies with poor quality or high risk of bias and to test the stability of the results of analyses. In addition, we will conduct cluster analysis of each outcome index to screen antiviral drugs with excellent efficacy and low side effects in the treatment of COVID-19.

### Publication bias

If more than 10 eligible studies are included, funnel plots[57] will be drawn and analyzed using State version 17 to visualize potential publication bias. If the points represented by included studies all clustered at the top of funnel plot and symmetrical, it indicates there is less possibility of publication bias. Moreover, we will use Begg’s and Egger’s methods[58, 59] to quantify potential publication bias and data statistics.

### Strength of evidence

The quality of evidence in studies shall be independently evaluated by two investigators based on the Grading of Recommendations Assessment, Development and Evaluation (GRADE) system[60], and the evidence strength will be shown as high, moderate, low or very low.

### Discussion

The COVID-19 pandemic has resulted in a global crisis in public health[61, 62], however, there are still no safe and effective drugs to resist COVID-19 until now. We will use network meta-analysis to analysis available evidence from RCTs to compare the safety and efficacy of four antiviral drugs (including Ribavirin, Arbidol, Chloroquine Phosphate and Interferon) alone or in combination, in patients with COVID-19 on the basis of standard treatment. At the same time, subgroup analysis and sensitivity analysis will be performed on the included studies to reveal the potential impact of RCTs quality on overall results. In addition, we shall use the GRADE method to evaluate the strength of studies evidence. This study is expected to provide high quality network meta-analysis, together with subgroup analysis, sensitivity analysis and publication bias analysis, to reveal four antiviral drugs alone or in combination to reveal the robustness and strength of evidence for relative efficacy against COVID-19, which will provide better evidence for future clinical decision-making.

### Limitations

Effective regimens for intervention of COVID-19 involves a combination of multiple departments and treatment protocols. In addition, the treatment options will vary among patients because of differences in patients’ basic characteristics as well as in adherence to treatment. The purpose of our study is mainly to evaluate efficacy and safety of four antiviral drugs against COVID-19 in guidelines, so the differences of other treatments on results will be not considered in study and not involve the difference of therapeutic effect caused by different doses of drugs, which will be an active area of investigation for our research group.

Second, the following shortcomings inevitable exist during literature collection: the quality of part studies not being high enough, A small number of patients being enrolled in part studies, literature excluded not published in English or Chinese, and the sample size of included not being large enough.

## Data Availability

No datasets were generated or analysed during the current study. All relevant data from this study will be made available upon study completion.

## Supporting information

S1 Checklist. PRISMA-P 2015 checklist. (DOCX)

## Author Contributions

**Conceptualization:** Liang-kun Zhang, Wen-chao Gu, and Ling Li. **Methodology:** Liang-kun Zhang, Wen-chao Gu, Ling Li, and Jian Chen. **Supervision:** Jian Chen.

**Software:** Liang-kun Zhang, Wen-chao Gu.

**Project administration:** Jian Chen.

**Writing - original draft:** Liang-kun Zhang, Wen-chao Gu.

**Writing - review & editing:** Liang-kun Zhang, Wen-chao Gu and Ling Li.

